# Psychological Impact of the Galleri Test (sIG(n)al): Protocol for a longitudinal evaluation of the psychological impact of receiving a cancer signal in the NHS-Galleri Trial

**DOI:** 10.1101/2023.06.12.23291276

**Authors:** Laura Marlow, Ninian Schmeising-Barnes, Jane Warwick, Jo Waller

**Affiliations:** King’s Cancer Prevention Group, School of Cancer and Pharmaceutical Sciences, King’s College London, London, UK

## Abstract

**Introduction:** Multi-cancer early detection (MCED) blood tests look for cancer signals in cell-free deoxyribonucleic acid (cfDNA). These tests have the potential to detect cancers at an earlier (asymptomatic) stage, improving cancer outcomes. Any screening method needs careful consideration of the psychological harms prior to implementation. The aim of this research is to explore the psychological impact of having a cancer signal detected following an MCED blood test.

**Methods and Analysis:** The project is embedded in the NHS-Galleri trial (ISRCTN91431511; NCT05611632), a large clinical trial that has randomised over 140,000 members of the general population aged 50-77 1:1 to either the intervention (blood tested with MCED test) or control (blood stored) arm. This work focuses on participants in the intervention arm of the trial who have a cancer signal detected. All participants who have a cancer signal detected (expected to be around 700 assuming a 1% test positive rate) will be sent a questionnaire at three time points: soon after receiving their result, 6-months and approximately 12-months later. The primary outcome is anxiety, assessed using the short-form State Trait Anxiety Inventory (STAI-6). We will also assess the psychological consequences of screening (using the Psychological Consequences of Screening Questionnaire), reassurance and concern about the test result, as well as understanding of results, cancer risk perceptions and help- and health-seeking behaviour. A sub-sample of 40 participants (20 with a cancer diagnosis and 20 for whom no cancer was found) will be invited to take part in a one-to-one semi-structured interview to explore their experience in depth.

**Ethics and Dissemination:** Ethical approval for this work has been granted by the Wales Research Ethics Committee as part of the NHS-Galleri trial (Ref 21/WA/0141). Results will be disseminated via peer-reviewed publication and presentations at national and international conferences.

**Strengths and limitations:**

- This will be the first UK study to explore the psychological impact of an MCED screening blood test.
- Multiple aspects of psychological impact will be considered across three time points ensuring our understanding of impact is wide ranging and extends beyond anxiety alone.
- Test results are only communicated to participants if a cancer signal is found (in order to maintain blinding), so it is not possible to collect data from those receiving a negative result following their first blood test. Comparative information on psychological impact will need to be made with previous research in the cancer screening context.
- The findings could be used to support policy making by the UK National Screening Committee (UKNSC) regarding recommendations for MCED screening in the UK in the future.
- Data will be collected within the context of a clinical trial, so our findings will need to be interpreted with appropriate caution.

## INTRODUCTION

Population-based cancer screening aims to identify signs of cancer among asymptomatic individuals at an early stage. The goal is to achieve better outcomes than would be expected if cancer was only identified following symptomatic presentation. The UK National Screening Committee (NSC) currently recommends population-based screening for cervical, breast and bowel cancer and all three programmes are estimated to have saved thousands of lives.^1^ In 2022, the UKNSC also recommended that targeted lung cancer screening be rolled out for people at high risk. A 2019 review of screening programmes in England stated that “Screening programmes are effectively judged on whether the benefits to those who get earlier treatment outweigh the harms to those people who get treated unnecessarily, or who are subject to unnecessary anxiety.” (p18)^2^ The premise of screening is such that most healthy individuals will not benefit from participation, so participation in screening involves risk-benefit considerations for all who take part.^3^

Most research evidence exploring psychological harms following screening focuses on anxiety as the primary endpoint.^4^ Anxiety is a mental state associated with intense emotion, worry or apprehension.^5^ People who receive ‘positive’ (sometimes called abnormal) cancer screening results can have higher anxiety levels than those who test ‘negative’ or are not tested, as demonstrated in the context of cervical screening (Kitchener, Burns et al. 2004, Posner, Vessey 1988),^6;7^ colorectal cancer screening,^8;9^ and mammography.^10-12^ For those receiving a positive screening result that does not ultimately lead to a diagnosis of cancer, or a pre-cancerous condition that requires treatment (sometimes called a ‘false positive’ result), raised anxiety is most evident in the short term, but can continue for a significant period afterwards, especially if diagnostic tests are invasive.^13^ Though anxiety can reduce over the months following the test, there is evidence that outcomes which are more test-specific, such as cancer worry and perceived risk of a future diagnosis can persist.^14;15^ ‘Positive’ screening results also have the potential to influence health-behaviours more broadly^16;17^ acting as a motivating or demotivating factor for future health behaviours and reattendance. Studies in existing screening contexts provide little evidence of false reassurance among those with false-positive results, but future research should continue to explore this as a possibility.^18^ Consequently, it is vital that the psychological harms and subsequent behavioural impacts are considered thoroughly before screening is offered at a population level. Evaluations of new screening modalities are therefore encouraged to include assessment of psychological harms.^19^

A novel approach to cancer screening is to use multi-cancer early detection (MCED) blood testing. MCED blood tests look for cancer ‘signals’ in cell-free deoxyribonucleic acid (cfDNA) and have the potential to identify multiple cancer types. There are currently several blood-based MCEDs in development,^20^ but evidence regarding their clinically relevant impact on cancer outcomes is yet to be determined. To be considered acceptable for population-wide screening, these tests should detect multiple cancer types, including cancers associated with lowest survival, accurately identify the tumour site, have low false-positive rates and high positive predictive values and ideally be affordable and cost-effective.^20^ Understanding the potential psychological impact of having a cancer signal detected is also a vital part of evidence needed alongside clinical outcomes.^21^

In 2021, a randomised controlled trial (RCT) trial began in England (Trial name: NHS-Galleri trial, ISRCTN91431511; NCT05611632). This trial is designed to assess whether offering a blood-based MCED test (Galleri® test, GRAIL, LLC.) for men and women aged 50-77 years, without personal history of invasive cancer within the last 3 years, can reduce the number of late-stage cancers diagnosed.^22^ Within the trial, participants are invited for three blood tests at 12-month intervals. Those with a cancer signal is detected are referred into an NHS urgent care referral pathway or rapid diagnostic pathway for diagnostic testing. Participants who do not have a cancer found following diagnostic testing are invited for second and third rounds of screening at 12 and 24 months. Our work on the psychological impact of having a cancer signal detected via MCED testing (Acronym: sIG(n)al) is nested within the NHS-Galleri trial. The aim is to assess the psychological impact of having a cancer signal detected at three time points; shortly after receiving MCED test results, and then at 6- and approximately 12-month follow-up.⍰ Our primary objects are i) to establish levels of anxiety among participants, who have been informed that a cancer signal was detected, shortly after receiving their results, ii) to compare longer-term anxiety between those subsequently diagnosed with cancer and those who have a diagnostic work-up but no cancer is found, at 6- and 12-month follow-up and iii) to explore in depth the experiences of men and women who have a cancer signal detected (using qualitative methods).

## METHODS AND ANALYSIS

### Design

A longitudinal observational design with a nested qualitative study. Survey data are collected at three time-points. Semi-structured one-to-one interviews will be carried out to explore patient experiences and understanding of results in more depth.

### Participants and Eligibility

The NHS-Galleri trial has enrolled over 140,000 participants aged 50-77 years (August 2021-July 2022) across eight participating cancer alliances, randomising to control or intervention arms (1:1). Participants in the intervention arm who have a cancer signal detected after their first MCED test are sent paper questionnaires at three time points unless they opt out (See Figure 1). A sub-sample of 40 participants (20 with a cancer diagnosis and 20 with a cancer signal detected but no cancer found following diagnostic work-up) are invited to take part in an interview to explore their experience in depth. In line with the trial protocol design,^22^ participants who do not have a cancer signal detected remain blinded to their allocation arm and are not given an explicit test result; we are thus unable to recruit a comparison group receiving a negative result.

### Procedures

Paper questionnaire packs are sent to participants’ homes. These include a participant information sheet explaining the purpose of the survey, a paper survey and a freepost envelope for returning completed questionnaires to the research team at King’s College London. An option to complete the questionnaire online is also available, by visiting a website printed on the information sheet and the front of the questionnaire. Participants need to enter a unique code (also printed on the questionnaire) to access the survey. Questionnaires are sent at three timepoints (see box 1 for details). All surveys are available on Open Science Framework (OSF): https://osf.io/uj86p.

Within the 6-month questionnaire, participants are asked to indicate if they are happy to be contacted about an interview. Interviewees who consent to be contacted are selected purposefully to represent a range of characteristics (age, gender, index of multiple deprivation [IMD; an area-level measure of relative deprivation based on Lower-layer Super Output Areas or neighbourhood]^23^), ethnicity and self-reported cancer diagnosis) based on their questionnaire data. Since recruitment for the interviews is over 10 months (mirroring the 10-month trial recruitment period), we will not have all participants opting into being interviewed at the same time and purposeful recruitment is thus an iterative process with ongoing review. Interviews are semi-structured and follow a topic guide (available on OSF: https://osf.io/cau6p). They are carried out by LM or NS (author initials), and are face-to-face or on the telephone, depending on the preference of the interviewee. Interviews last up to 1-hour and participants are given a £40 voucher to compensate them for their time. All interviews are audio-recorded and transcribed verbatim for analysis. The researchers also keep reflexive journals throughout the process of data collection and during analysis of the data.

### Primary outcomes

Our main primary outcome measure is anxiety assessed using the short-form version of the Spielberger State-Trait Anxiety Inventory (STAI-6).^24;25^ The STAI is a measure of state anxiety that has been validated and used in many studies and is the most commonly used measure of anxiety in the context of cancer screening.^4^ We will therefore be able to compare our findings to those from other relevant studies in the UK (e.g. in HPV Primary screening^26^) and worldwide (e.g. in lung screening^27^). We may also be able to compare the results of our study with those that have used the Hospital Anxiety and Depression Scale (HADS-A) using suggested equivalence scores.^28^ The STAI also has cut-offs indicative of ‘high anxiety’ that might be expected to lead to a clinical diagnosis, though the exact cut-offs used do vary across studies.^4^

Despite the benefits of using the STAI to assess anxiety, some researchers have suggested that this measure is not sensitive enough or sufficient for identifying changes in psychological outcomes following positive screening results.^29^ To address this, we will include additional measures of psychological impact (see Table 1). The first is the Psychological Consequences of Screening questionnaire (PCQ) which measures the impact of screening on an individual’s emotional, social, and physical functioning.^30^ The PCQ also offers the opportunity to assess positive longer-term psychological impact following screening. We will also assess result-specific concern and reassurance, as studies have shown that result-specific outcomes (e.g. concern about the test result or future risk of cancer) are more likely to persist longer term.^31;32^ In addition to the measures of psychological impact, two types of behavioural impact will be considered as primary outcome measures. These apply exclusively to participants who have a cancer signal detected at the first blood test but no cancer is found. The first is self-reported behaviour change in relation to routine cancer screening attendance and symptomatic help-seeking (reported in the T3 survey). The second is reattendance for a second trial blood test (at 12 months).

**Table 1:** Summary of primary and secondary outcome variables.

| Construct | Details | Validated scale | T1 | T2 | T3 |  |  |
| --- | --- | --- | --- | --- | --- | --- | --- |
|  |  |  |  |  | a | b | C |
| **Primary outcomes** |  |  |  |  |  |  |  |
| Anxiety | STAI-6<sup>25</sup> (6-items, 4-point likert scale) | ? | ? | ? | ? | ? | ? |
| Emotional, physical and social consequences of screening | PCQ<sup>30</sup> (negative: 12-items, 4-point likert scale) | ? | ? | ? | ? | ? | ? |
|  | PCQ<sup>30</sup> (positive: 10-items) | ? |  | ? | ? | ? |  |
| Test-specific concern | Single-item (5-point likert) |  | ? |  | ? |  |  |
| Test specific reassurance | Single-item (5-point likert) |  | ? | ? | ? |  |  |
| Changes in health behaviour | Single item for each behaviour |  |  |  | ? |  |  |
| **Secondary outcomes** |  |  |  |  |  |  |  |
| Understanding of results | Understanding of result meaning (1-item, 6 response options) | ? | ? |  | ? | ? | ? |
|  | Confidence in understanding (1-item, 5-point likert scale) |  | ? | ? | ? |  |  |
| Perceived risk of cancer | Affective risk (1-item, 5-point likert scale) |  | ✓ | ✓ | ✓ |  |  |
|  | Deliberative risk (1-item, 5-point likert scale) |  | ✓ | ✓ | ✓ |  |  |
| Overall experience | 1-item, 4 response options |  | ? |  |  |  |  |
| Satisfaction with the trial | Multiple items assessing satisfaction (5-point likert scale): |  |  |  |  |  |  |
|  | Invitation; information; plan for follow-up |  | ? |  |  |  |  |
|  | Appointment, result delivery; |  | ? |  | ? |  |  |
|  | Consultations; Support during follow-up |  |  | ? |  |  |  |
| Future intention to have a Galleri test | Single-item |  | ✓ | ✓ | ✓ | ✓ | ✓ |
| Decisional Regret | Decisional Regret Scale<sup>33</sup> (5-items, 5-point likert scale) | ✓ |  |  | ✓ | ✓ | ✓ |
| Fear of recurrence | Fear of Cancer Recurrence-4<sup>34</sup> (4-items, 5-point likert scale) | ✓ |  |  |  | ✓ |  |

T1 (Time 1); T2 (Time 2); T3 (Time 3). a: returning for second blood draw; b: with cancer found; c: Non-attenders.

### Secondary outcomes

Secondary outcomes assess additional constructs relevant to psychological impact (see Table 1) including understanding of the test result, perceived cancer risk and satisfaction with different aspects of the trial, future intention to have the Galleri test and decisional regret (using the Decision Regret Scale^33^). We will also measure fear of cancer recurrence among those diagnosed with cancer (using the Fear of Cancer Recurrence-4^34^).

### Participant characteristics and explanatory variables

Sociodemographic variables including age, IMD, biological sex, ethnicity, country of birth, marital status and education are collected at the first clinic visit following consent. Additional variables assessing cognitions, emotions and behaviours (assessed in the three surveys) will be treated as explanatory variables. These constructs are expected to facilitate understanding of differences in psychological outcomes and include coping and appraisal, cancer worry and attitudes to cancer and early detection.

### Sample Size

The maximum available sample size is determined by expected numbers recruited in the NHS-Galleri trial – questionnaires are sent to all those with a cancer signal detected. Approximately 1% of the intervention arm (n∼700) are expected to have a cancer signal detected. We anticipate a 50% response rate at each time point, resulting in data from up to 350 participants at each timepoint (if positivity rates are as expected). This sample size will allow us to estimate the proportion of participants experiencing ‘very high’ anxiety (using a STAI score >49 consistent with^26^) with a precision of +/- 3 to 4% (based on 10-20% experiencing ‘very high’ anxiety). The anticipated sample size would also give us 88% power (alpha=.05) to detect a mean difference in STAI-score of 4-points (SD=12) between those who go on to have a cancer diagnosed and those for whom no cancer is found following further tests. The decision to conduct 40 interviews is predominantly pragmatic, taking into consideration the expected information power of the sample.^35^

### Data Analysis and statistics

Survey data will be analysed in SPSS and STATA. Findings will be considered statistically significant where p<0.05, however we acknowledge that the use of p-values to indicate significance has its limitations. Confidence intervals will also be presented. If there are significant demographic differences between participants who do and do not respond to the questionnaire by sex or IMD, we will calculate and apply weights to adjust for the possibility that the sample may not represent those with a cancer signal detected in the trial in relation to sex and deprivation level. All analyses will adjust for recruitment location (i.e. one of eight cancer alliances). Descriptive statistics will be reported for all primary outcomes and secondary outcomes at each timepoint. This will include anxiety (STAI score and % with ‘very high’ anxiety), psychological consequences of screening (total score and three separate dimensions: emotional, physical and social), test-specific concern and reassurance, understanding of results, information seeking and satisfaction. Mean STAI and PCQ scores (with SDs) will be reported by sociodemographic and other explanatory variables. Hierarchical linear regression will be used to explore relationships between explanatory variables and continuous outcomes. Similarly, stepwise logistic regression will be used to explore relationships between explanatory variables and categorical outcomes. Profile analysis will be used to explore whether combined responses (regardless of cancer diagnosis) are different at any particular point in time i.e. by testing the “flatness” hypothesis over time (i.e. T1, T2, T3) for each outcome. The Hotelling’s Trace F(df), alpha and p-value will be reported. Statistical differences between those with and without cancer diagnosed will be explored using profile analysis to establish whether one result group, on average, scores differently on STAI and PCQ scores regardless of time-point (the “parallelism” hypothesis) and also whether cancer patients and those with no cancer found following further tests have a similar pattern of response over the course of time (the “levels” hypothesis).

Qualitative data collected during the semi-structured interviews will be analysed using reflexive thematic analysis. This is an interpretive approach to analysing qualitative data supporting the researcher to identify themes and patterns in the data set.^36^ Reflexive TA recognises that themes are actively created by the researcher at the intersection of data and interpretive engagement, rather than awaiting discovering. There are six phases of Reflexive TA: familiarisation, coding, generating themes, reviewing and developing themes, refining and defining themes and producing the report. These can overlap and there is flexibility as the researchers move through these phases. The software NVIVO will be used to facilitate data coding and management.

### Patient and public involvement (PPI)

A PPI Panel was established in May 2021. Our panel includes five representatives aged 50-70 years old and includes people with and without personal cancer experience. Our PPI panel has supported the design and the development of the study protocol and study materials (participant information sheets, consent forms, questionnaires) and will contribute to patient/public facing dissemination. Field notes are used to record all discussions. Contributions to the materials are documented and we will make these available in a working paper on OSF. PPI activity will be reported using the GRIPP2 checklist^37^ to ensure quality and transparency.

## DISCUSSION

The findings will help to improve understanding of the psychological impact of having a cancer signal detected by MCED screening and inform the development of procedures, supporting information and interventions to minimise MCED screening-associated anxiety. Findings will inform UK National Screening Committee (UKNSC) recommendations regarding adoption of MCED screening and will support any future roll-out. We will also be able to explore whether psychological impact is pronounced in particular groups (e.g. by age or biological sex), and whether particular population sub-groups may consequently require additional support if MCED testing is implemented at a national level.

Comparisons will be made with previous research in the cancer screening context. Preliminary data from the prospective Pathfinder study,^38^ in the US suggest that having a cancer signal detected through MCED screening can result in increased anxiety, relative to those that do not have a cancer signal found, but this decreased towards baseline within 12-months. This will be the first UK study to explore anxiety and other psychological outcomes in the context of MCED blood testing within an asymptomatic population. Exploring this within the trial context means that psychological outcomes can contribute to decisions about implementation, rather than being an afterthought. This is an important step in acknowledging the potential for screening to result in a range of harms, including non-physical harms.^39^

Considerable steps have been taken in the context of the NHS-Galleri trial to ensure that the recruited sample represents the wider population in terms of age and deprivation and that sufficient numbers are recruited from groups typically under-represented in clinical trials (e.g. those from ethnic minority backgrounds). However, there may still be some volunteer bias particularly in the sIG(n)al study where participation requires completion of additional questionnaires, as opposed to just clinical data.

Logistical aspects of delivery (e.g. how and where blood test appointments are available and how results are communicated) have the potential to influence experiences and consequently emotional responses. The way that the test has been offered in the clinical trial context may not reflect exactly how an MCED screening programme would look if implemented, so the sIG(n)al findings may not be fully generalisable to a routine context.

The trial design has also influenced the research questions for the sIG(n)al sub-study. For example, findings will allow us to explore the psychological impact of particular result combinations offered within repeated MCED screening (including those who have a signal detected again in round 2 screening and those that no longer have a signal detected). However, because results are not communicated to participants who do not have a cancer signal detected in round 1 (to maintain trial blinding), we will not be able to explore the impact on those who only have a signal detected at the second screening round.

The sIG(n)al sub-study has been designed to provide a holistic understanding of the broad psychological impact of MCED screening, as well as including qualitative work to explore experiences in more depth. The limited timeline for the project makes it difficult to ascertain longer term impact on behaviour, but we will assess self-reported changes to inform further investigation of this.

## Data Availability

N/A - the article reports a protocol.

## ETHICS AND DISSEMINATION

Ethical approval for this work has been granted by the Wales Research Ethics Committee as part of the NHS-Galleri trial ethics application (Ref 21/WA/0141). The studies have also been registered on the King’s Data Protection Register (ID#17436). Results will be disseminated via peer-reviewed publication and presentations at national and international conferences. Reporting will be supported by the STROBE^40^ and COREQ guidelines.^41^

## Author Contributions

Jo Waller and Laura Marlow conceived of the study. Laura Marlow and Jo Waller developed the protocol, with statistical support from Jane Warwick. Laura Marlow, Jo Waller and Ninian Schmeising-Barnes developed the study measures. Laura Marlow drafted the paper. All authors contributed to the final version of the manuscript. The GRAIL, LLC publications team reviewed the final manuscript prior to submission.

## Acknowledgements

The sIG(n)AL Sub Study is run by the KCL Cancer Prevention Group (CPG) Behavioural Science Team. The Behavioural Science Team have worked in close collaboration with the CPTU NHS Galleri Project Team, but are a separate, autonomous group. We would like to thank our PPI contributors for their collaboration on this project.

## Box 1

Description of the surveys (all surveys are available at https://osf.io/uj86p).

|  |  |
| --- | --- |
| Time 1 (T1) | Sent to all participants with a cancer signal found along with their written results letters, approximately 7-days after they have been given their results by a research nurse over the phone. |
| Time 2 (T2) | Sent to participants with a cancer signal found 6-months after their written results letter was dispatched. A single survey is sent, but sections of the survey are designed for 'If you were told you have cancer' AND 'If no cancer was found following tests'. |
| Time 3 (T3) | Sent approximately 12-14 months after the initial result. Three slightly different versions of T3 are used as follows:<br>a. Participants who attend a second blood test.<br>b. Participants identified as having received a cancer diagnosis and undergoing treatment (and so not offered a second blood test within the trial).<br>c. Participants who do not attend their second blood test appointment.<br>Differences in the content of these versions are outlined in Table 1. |

Note: All surveys are available at https://osf.io/uj86p.

